# The prognostic value of lymph node involvement after neoadjuvant chemotherapy is different among breast cancer subtypes

**DOI:** 10.1101/2020.12.05.20244582

**Authors:** Lucie Laot, Enora Laas, Noemie Girard, Elise Dumas, Eric Daoud, Beatriz Grandal, Jean-Yves Pierga, Florence Coussy, Youlia Kirova, Elsy el Alam, Guillaume Bataillon, Marick Lae, Florence Llouquet, Fabien Reyal, Anne-Sophie Hamy

**Author notes:** **Corresponding author:** Pr Fabien REYAL, Institut Curie, Department of Surgery, 26 rue d’Ulm, 75005 Paris, 00 33 615271980. Contributed equally (Lucie Laot, Enora Laas).

## Abstract

**Introduction:** The three different breast cancer subtypes (Luminal, *HER2-*positive and triple negative (TNBCs) display different natural history and sensitivity to treatment, but little is known about whether residual axillary disease after neoadjuvant chemotherapy (NAC) carries a different prognostic value by BC subtype.

**Methods:** We retrospectively evaluated axillary involvement (0, 1 to 3 positive nodes, ≥ 4 positive nodes) on surgical specimens from a cohort of T1-T3NxM0 BC patients treated with NAC between 2002 and 2012. We analyzed the association between nodal involvement (ypN) binned into 3 classes (0; [1-3];4 or more), relapse-free survival (RFS) and overall survival (OS) among the global population, and according to BC subtypes.

**Results:** 1197 patients were included in the analysis (luminal (n = 526, 43.9%), TNBCs (n = 376, 31.4%), *HER2*-positive BCs (n = 295, 24.6%)). After a median follow-up of 110.5 months, ypN was significantly associated with RFS, but this effect was different by BC subtype (P_interaction_= 0.004), and this effect was nonlinear. In the luminal subgroup, RFS was impaired in patients with 4 or more nodes involved (HR=2.8; 95% CI [1.93;4.06], *p*<0.001) when compared with ypN0, while it was not in patients with 1 to 3 nodes (HR=1.24, 95% CI = [0,86;1.79]). In patients with TNBC, both 1-3N+ and ≥ 4 N+ classes were associated with a decreased RFS (HR=3.19, 95%CI= [2.05; 4.98] and HR=4.83, 95%CI= [3.06; 7.63], respectively *versus* ypN0, *p*< 0.001). Similar decreased prognosis were observed among patients with *HER2*-positive BC (1-3N+: HR=2.7, 95%CI= [1.64; 4.43] and ≥ 4 N+: HR=2.69, 95%CI= [1.24; 5.8] respectively, *p*=0.003).

**Conclusion:** The prognostic value of residual axillary disease should be considered differently in the 3 BC subtypes to accurately stratify patients with a high risk of recurrence after NAC who should be offered second line therapies.

## Introduction

Neoadjuvant chemotherapy (NAC) has been for decades the cornerstone of treatment strategy for locally advanced breast cancers (BC) (T3-T4), and tumors not accessible to conservative treatment. Since the publication of the CREATE-X (1) and the KATHERINE trial (2), it also became a standard of care in triple negative (TNBCs) and *HER2*-positive BC. Beyond the increase of breast conservative surgery rates, NAC provides a way to assess tumor chemosensitivity and evaluate mechanisms of resistance to chemotherapy through the evaluation of residual tumor burden.

Axillary lymph node involvement is the most important prognostic factor in BC, and has long been proven to be correlated with poor survival outcomes (3) (4) (5) (6). In the neoadjuvant setting, several studies have established the critical role of nodal burden in the assessment of prognosis after NAC in large cohorts of patients (7) (8) (9) (10)(11).

Pathologic complete response (pCR) is defined as the absence of invasive cancer in the breast and axillary lymph nodes, and has been shown to be associated with a better long-term survival among BC patients treated with NAC. Although nodal axillary response has been described as a superior prognostic parameter after NAC (12) (13), overall pCR is more frequently used and has been adopted by the Food and Drug Administration and the European Medicines Agency as an important endpoint in BC neoadjuvant studies (14).

The prognostic value of pCR to predict event-free survival varies among BC subtypes(15) (16). In 2014, a meta-analysis by Cortazar *et al*. (17) including 11 955 patients found a stronger association between pCR and long-term outcomes in patients with TNBCs (RFS: HR=0.24, 95% CI [0.18-0.33]) and in those with *HER2*-positive hormone receptor negative BC (RFS: HR=0.15, 95% CI [0.09-0.27]); whereas the association was less marked in *HER2*- positive hormone receptor positive BC (RFS: HR=0.58, 95% CI [0.42-0.82]) and luminal BC (RFS: HR=0.49, 95% CI [0.33-0.71]).

However, the evidence evaluating the prognostic impact of residual axillary burden after NAC according to BC subtypes is scarce. Most of the studies evaluating the prognostic impact of axillary response to NAC classified patients in a binary manner - depending on the presence or absence of residual nodal disease - without taking into account the number of axillary lymph nodes involved – and few studies - if any - performed upfront comparison of the prognostic significance by BC subtype.

The aim of our study was to evaluate the impact of the number of axillary nodes involved on survival outcomes according to BC subtype in a real-life cohort of breast cancer patients treated with NAC.

## Material and methods

### Patients

We analyzed a previously described retrospective cohort of patients (18) (19) with invasive breast carcinoma stage T1-T3NxM0 and treated with NAC at Institut Curie, Paris, between 2002 and 2012 (NEOREP Cohort, CNIL declaration number 1547270). We included unilateral, non-recurrent, non-inflammatory, non-metastatic tumors, excluding T4 tumors. All patients received NAC, followed by surgery and radiotherapy. NAC regimens changed over our recruitment period (anthracycline-based regimen or sequential anthracycline-taxanes regimen), with trastuzumab used in an adjuvant and/or neoadjuvant setting since 2005. Endocrine therapy (tamoxifen or aromatase inhibitor) was prescribed when indicated. The study was approved by the Breast Cancer Study Group of Institut Curie and was conducted according to institutional and ethical rules concerning research on tissue specimens and patients. Informed consent from patients was not required by French regulations.

### Tumor samples and pathological review

#### BC subtypes

Cases were considered estrogen receptor (ER) or progesterone receptor (PR) positive (+) if at least 10% of the tumor cells expressed estrogen and/or progesterone receptors (ER/PR), in accordance with guidelines used in France (20). HER2 expression was determined by immunohistochemistry with scoring in accordance with American Society of Clinical Oncology (ASCO)/College of American Pathologists (CAP) guidelines (21). Scores 3+ were reported as positive, score 1+/0 as negative (-). Tumors with scores 2+ were further tested by FISH. HER2 gene amplification was defined in accordance with ASCO/CAP guidelines. We evaluated a mean of 40 tumor cells per sample and the mean HER2 signals per nuclei was calculated: a HER2/CEN17 ratio ≥ 2 was considered positive, and a ratio < 2 negative. BC subtypes were defined as follows: tumors positive for either ER or PR, and negative for HER2 were classified as luminal; tumors positive for *HER2* were considered to be *HER2*-positive BC; tumors negative for ER, PR, and *HER2* were considered to be triple-negative breast cancers (TNBC). Tumor cellularity was defined as the percentage of tumor cells (in situ and invasive) on the specimen (biopsy or surgical specimen). Mitotic index was reported per 10 high power fields (HPF) (1 HPF= 0.301 mm2).

#### Post-NAC nodal involvement (ypN)

Post-NAC nodal involvement (ypN) was divided into three categories: no axillary involvement (ypN = 0), intermediate involvement (1 to 3 nodes involved, 1 ≤ ypN ≤ 3) and high axillary involvement (4 or more nodes involved, ypN ≥4). Nodal extent was also analysed as a continuous variable.

#### Residual Cancer Burden index (RCB)

Histological components of the “Residual Cancer Burden” were retrieved for calculating the score as described in 2007 by Symmans (22). RCB index enables the classification of residual disease into four categories: RCB-0 (complete pathologic response = pCR), RCB-I (minimal residual disease), RCB-II (moderate residual disease) and RCB-III (extensive residual disease). RCB was calculated through the web-based calculator that is freely available on the internet (www.mdanderson.org/breastcancer_RCB).

#### TILs and LVI

Lymphovascular invasion (LVI) was defined as the presence of carcinoma cells within a finite endothelial-lined space (a lymphatic or blood vessel). Tumor infiltrating lymphocytes (TILs) were defined as the presence of mononuclear cells infiltrate (including lymphocytes and plasma cells, excluding polymorphonuclear leukocytes), and were also evaluated retrospectively for research purposes, according to the recommendations of the international TILs Working Group (23), (24).

### Study endpoints

Relapse-free survival (RFS) was defined as the time from surgery to death, loco-regional recurrence or distant recurrence, whichever occurred first, and overall survival (OS) was defined as the time from surgery to death. Patients for whom none of these events were recorded were censored at the date of their last known contact. Survival cutoff date analysis was 1 February 2019.

### Statistical analysis

The study population was described in terms of frequencies for qualitative variables, or medians and associated ranges for quantitative variables. Chi-square tests were performed to search for differences between subgroups for each variable (considered significant for p- values ≤ 0.05). Survival probabilities were estimated by the Kaplan–Meier method, and survival curves were compared in log-rank tests. Hazard ratios and their 95% confidence intervals were calculated with the Cox proportional hazards model. Variables with a *p*-value for the likelihood ratio test equal to 0.05 or lower in univariate analysis were selected for inclusion in the multivariate analysis. A forward stepwise selection procedure was used to establish the final multivariate model and the significance threshold was 5%.

#### Linearity tests

For representations of the relationships between the nodal extent as a quantitative variable and the RFS and OS, we modeled these variables with cubic splines or polynomials with an order of more than one, before inclusion in the survival models, respectively. To assess the linearity of the relationship, by determining the deviation of the model from a straight line. We retained the model with the lowest AIC. Data were processed and statistical analyses were carried out with R software version 3.1.2 (www.cran.r-project.org, (R Foundation for Statistical Computing, 2009).

## Results

### Baseline patients’ and tumors’ characteristics

1197 patients were included in the cohort. Patients’ baseline characteristics are summarized in Table 1. Median age was 48 years old. Patient’s repartition by subtype was as follows: luminal (n=526, 43.9%), TNBC (n=376, 31.4%), *HER2*-positive (n=295; 24.6%).

**Table 1:** Patients and tumor characteristics by post-NAC nodal involvement.

| Characteristics | Class | All | Node negative | Node positive | *p* |
| --- | --- | --- | --- | --- | --- |
| **n =** |  | **1197 (100%)** | **682 (57%)** | **515 (43%)** |  |
| **Age median** |  | 48.5 (10.1) | 48.0 (10.4) | 49.3 (9.6) | 0.027 |
| **Age class** | [0 -50) | 678 (56.6) | 399 (58.5) | 279 (54.2) | 0.279 |
|  | [50 -60) | 352 (29.4) | 189 (27.7) | 163 (31.7) |  |
|  | 60+ | 167 (14.0) | 94 (13.8) | 73 (14.2) |  |
| **Menopausal status** | Premenopausal | 746 (62.8) | 432 (63.9) | 314 (61.3) | 0.396 |
|  | Postmenopausal | 442 (37.2) | 244 (36.1) | 198 (38.7) |  |
| **BMI** | 18.5-24.9 | 680 (57.1) | 401 (59.1) | 279 (54.5) | 0.302 |
|  | <18.5 | 48 (4.0) | 26 (3.8) | 22 (4.3) |  |
|  | 25-29.9 | 304 (25.5) | 160 (23.6) | 144 (28.1) |  |
|  | >=30 | 159 (13.4) | 92 (13.5) | 67 (13.1) |  |
| **Smoking status** | No | 719 (75.6) | 428 (76.4) | 291 (74.4) | 0.528 |
|  | Yes | 232 (24.4) | 132 (23.6) | 100 (25.6) |  |
| **BRCA mutation genes** | BRCA1 | 31 (11.7) | 24 (13.7) | 7 (7.8) | 0.404 |
|  | BRCA2 | 14 (5.3) | 10 (5.7) | 4 (4.4) |  |
|  | others | 1 (0.4) | 1 (0.6) | 0 (0.0) |  |
|  | No | 219 (82.6) | 140 (80.0) | 79 (87.8) |  |
| **Clinical T stage (TNM)** | T0-T1 | 70 (5.9) | 41 (6.0) | 29 (5.6) | **0.001** |
|  | T2 | 797 (66.6) | 483 (70.8) | 314 (61.1) |  |
|  | T3-T4 | 329 (27.5) | 158 (23.2) | 171 (33.3) |  |
| **Clinical N stage (TNM)** | N0 | 525 (43.9) | 372 (54.5) | 153 (29.8) | **<0.001** |
|  | N1-N2-N3 | 671 (56.1) | 310 (45.5) | 361 (70.2) |  |
| **BC subtype** | Luminal | 526 (43.9) | 197 (28.9) | 329 (63.9) | **<0.001** |
|  | TNBC | 376 (31.4) | 280 (41.1) | 96 (18.6) |  |
|  | HER2+ | 295 (24.6) | 205 (30.1) | 90 (17.5) |  |
| **ER status** | Negative | 544 (45.4) | 399 (58.5) | 145 (28.2) | **<0.001** |
|  | Positive | 653 (54.6) | 283 (41.5) | 370 (71.8) |  |
| **PR status** | Negative | 680 (57.9) | 450 (66.8) | 230 (46.0) | **<0.001** |
|  | Positive | 494 (42.1) | 224 (33.2) | 270 (54.0) |  |
| **Her2 status** | Negative | 902 (75.4) | 477 (69.9) | 425 (82.5) | **<0.001** |
|  | Positive | 295 (24.6) | 205 (30.1) | 90 (17.5) |  |
| **Histological type** | NST | 1060 (93.5) | 617 (96.0) | 443 (90.2) | **<0.001** |
|  | Others | 74 (6.5) | 26 (4.0) | 48 (9.8) |  |
| **KI67** | [0-10) | 65 (11.2) | 25 (7.9) | 40 (15.0) | **0.001** |
|  | [10-20) | 110 (18.9) | 49 (15.6) | 61 (22.9) |  |
|  | >=20 | 406 (69.9) | 241 (76.5) | 165 (62.0) |  |
| **Mitotic index** |  | 20.8 (19.2) | 24.1 (20.2) | 16.3 (16.9) | **<0.001** |
|  |  | 61.3 (18.2) | 61.0 (19.2) | 61.8 (16.4) |  |
| **SBR grade** | Grade I-II | 477 (41.3) | 200 (30.4) | 277 (55.7) | **<0.001** |
|  | Grade III | 678 (58.7) | 458 (69.6) | 220 (44.3) |  |
| **LVI** | No | 267 (61.0) | 158 (69.9) | 109 (51.4) | **<0.001** |
|  | Yes | 171 (39.0) | 68 (30.1) | 103 (48.6) |  |
| **DCIS component** | No | 601 (60.7) | 385 (66.3) | 216 (52.8) | **<0.001** |
|  | Yes | 389 (39.3) | 196 (33.7) | 193 (47.2) |  |
| **Stromal TIL levels (%)** |  | 24.0 (19.9) | 26.9 (21.7) | 19.3 (15.6) | **<0.001** |
| **IT TIL levels (%)** |  | 11.2 (12.3) | 12.4 (13.1) | 9.3 (10.6) | **0.001** |
| **CT regimen (NAC)** | anthra-taxanes | 841 (70.6) | 507 (74.7) | 334 (65.1) | **<0.001** |
|  | anthra | 235 (19.7) | 105 (15.5) | 130 (25.3) |  |
|  | taxanes | 25 (2.1) | 13 (1.9) | 12 (2.3) |  |
|  | others | 91 (7.6) | 54 (8.0) | 37 (7.2) |  |

Missing data: Menopausal status, n=9; BMI, n=6; Smoking status, n=246; BRCA mutation genes, n=932; Clinical T stage (TNM), n=1; Clinical N stage (TNM), n=1; PR status, n=23; Histological type, n=63; KI67, n=616; Mitotic index, n=117; NA, n=484; SBR grade, n=42; LVI, n=759; DCIS component, n=207; Stromal TIL levels (%), n=483; IT TIL levels (%), n=483; CT regimen (NAC), n=5'. The "n" denotes the number of patients. In case of categorical variables, percentages are expressed between brackets. In case of continuous variables, mean value is reported, with standard deviation between brackets. In case of nonnormal continuous variables, median value is reported, with interquartile range between brackets.

After NAC, 43% of the patients (515/1197) had a nodal involvement. Patients with bigger tumors, with clinical baseline nodal involvement, luminal BCs (*versus* TNBC or *HER2-* positive), low proliferative tumors (*versus* high proliferative), with lower immune infiltration (*versus* high TIL levels) were more likely to have a nodal involvement at NAC completion. The number of nodes ranged from 1 to 35 (median: 11) (Fig1A) and the number of lymph nodes involved varied from 0 to 21(Fig1B). In case of nodal involvement, the median number of nodes involved was 2 (Fig1C), and the repartition was significantly different among BC subtypes. Overall, 57% of the patients had no nodal involvement at axillar surgery (n=682), 28% had a mild nodal involvement (n=341), and 15% (n=174) had a high nodal involvement (Fig1D). This repartition was significantly different by BC subtype (*p*<0.001) (Fig1E).

**Figure 1:**
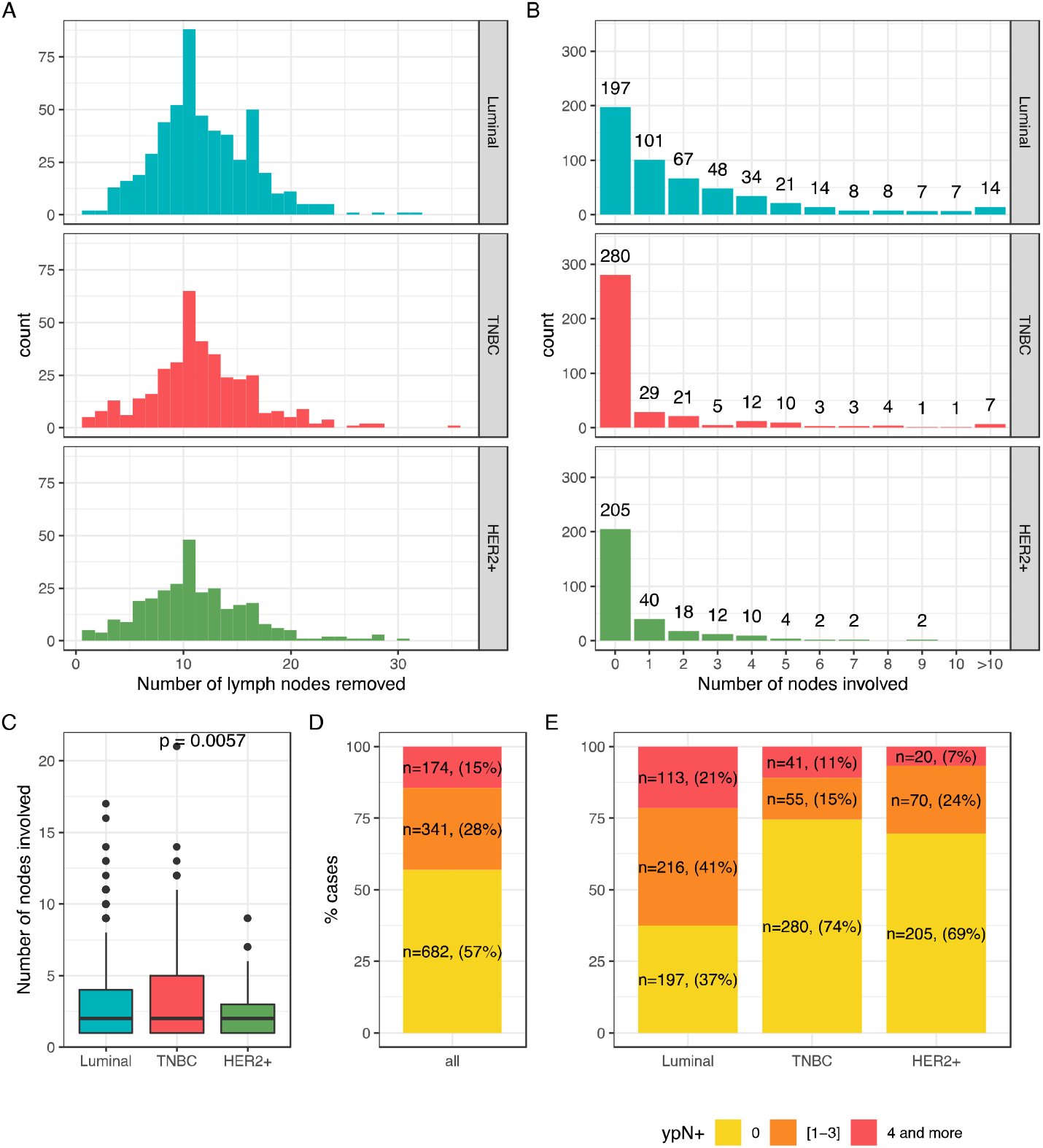
Nodal burden after NAC: Number of lymph nodes removed according to BC subtype (A), Number of involved nodes according to BC subtype (B); Mean number of nodes involved after NAC according to BC subtype (C), Node involvement repartition after NAC in the whole population (D) and according to BC subtype (E).

### Association between post-NAC involvement and tumor characteristics

Among post-NAC characteristics, node positivity was associated with RCB index (Table2, Fig2A), with the presence of lymphovascular invasion (Fig2B), and with higher post-NAC tumor cellularity (Fig2C). Neither post-NAC mitotic index (Fig2D), stromal (Fig2E) nor IT TILs (Fig2F) were significantly associated with post-NAC nodal status. Similar patterns were observed within each BC subtype (Figs S1A-F), with the very exception of post-NAC tumor cellularity (all 3 BC subtypes), post-NAC mitotic index (luminal BC), and str TILs levels (*HER2*-positive BC) that were significantly higher with increasing number of nodes involved (Figure S1).

**Table 2:** Tumor characteristics by post-NAC nodal involvement.

| Characteristics | Class | Post-NAC node involvement (ypN) |  |  | *p* |
| --- | --- | --- | --- | --- | --- |
|  |  | 0 | [1-3] | 4 and more |  |
| **n =** |  | 682 | 341 | 174 |  |
| **Pathological complete response** | No pCR | 396 (58.3) | 340 (100.0) | 173 (100.0) | **<0.001** |
|  | pCR | 283 (41.7) | 0 (0.0) | 0 (0.0) |  |
| **Post-NAC LVI** | No | 318 (78.3) | 156 (55.5) | 56 (38.6) | **<0.001** |
|  | Yes | 88 (21.7) | 125 (44.5) | 89 (61.4) |  |
| **RCB index (continuous)** |  | 1.0 (0.9) | 3.0 (0.8) | 3.6 (0.7) | **<0.001** |
| **RCB class** | RCB-0 | 200 (45.1) | 0 (0.0) | 0 (0.0) | **<0.001** |
|  | RCB-I | 53 (12.0) | 12 (6.4) | 0 (0.0) |  |
|  | RCB-II | 188 (42.4) | 101 (53.7) | 20 (24.1) |  |
|  | RCB-III | 2 (0.5) | 75 (39.9) | 63 (75.9) |  |
| **Stromal TIL levels (%) (post-NAC)** |  | 12.8 (13.1) | 13.6 (12.3) | 12.6 (12.1) | 0.750 |
| **IT TIL levels (%) (post-NAC)** |  | 7.2 (8.2) | 6.9 (8.0) | 5.7 (5.4) | 0.289 |
| **Mitotic index (post-NAC)** |  | 18.9 (30.9) | 12.6 (23.0) | 16.9 (34.8) | 0.103 |
| **Tumor cellularity (post-NAC)** |  | 19.6 (26.7) | 35.9 (25.5) | 36.5 (24.5) | **<0.001** |

Missing data: Pathological complete response, n=5; Post-NAC LVI, n=365; RCB index (continuous), n=483; RCB class: 0 ; RCB-0 ; ]0;1.36] : RCB-I ; ]1.36-3.28] : RCB-II ; >= 3.28 : RCB-III, n=483; Stromal TIL levels (%) (post-NAC), n=483; IT TIL levels (%) (post-NAC), n=715; Mitotic index (post-NAC), n=722; Tumor cellularity (post-NAC), n=483
1 The “n” denotes the number of patients. In case of categorical variables, percentages are expressed between brackets. In case of continuous variables, mean value is reported, with standard deviation between brackets. In case of nonnormal continuous variables, median value is reported, with interquartile range between brackets.

**Figure 2:**
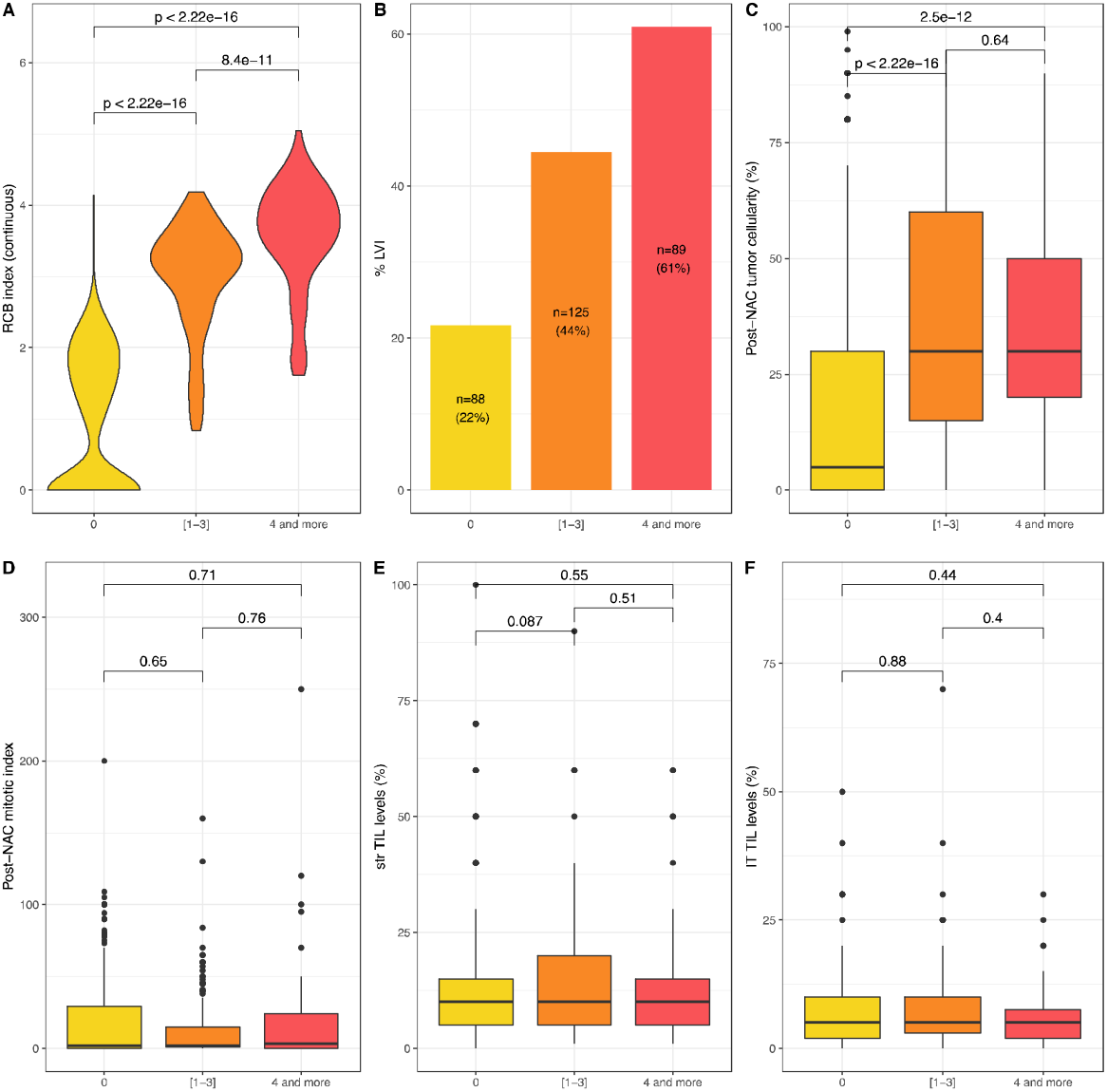
Association between post-NAC involvement and tumor characteristics in the whole population: RCB index (A); Lymphovascular invasion (B); Tumor cellularity (C); Post-NAC mitotic index (D); Stromal TIL levels (E). Intra tumoral (IT) TIL levels (F).

### Survival Analyses

With a median follow-up of 110.5 months, 371 patients experienced relapse, and 228 died. After univariate analysis, post-NAC nodal involvement was significantly associated with RFS in the whole population (*p*<0.001) (Table 3). After analyses by BC subtype, the association between nodal involvement binned by 3 classes and RFS was significant in all the BC subgroups, but this association was significantly different according to the BC subtype (*P*_interaction_ = 0.004). In the whole population, mild post-NAC nodal involvement (1 to 3); and high nodal involvement were associated with an impaired RFS (HR = 1.79, 95%CI [1.41 - 2.28] and HR=3.3, 95%CI [2.56 - 4.27]) (Fig3A).

**Table 3:** Association of clinical and pathological pre and post-NAC parameters with relapse-free survival after univariate and multivariate analysis in the whole population.

|  |  |  |  | Univariate |  |  |  | Multivariate |  |  |
| --- | --- | --- | --- | --- | --- | --- | --- | --- | --- | --- |
| Variable | Category | n | events | HR | 95% CI | p* | p | HR | 95% CI | p |
| Pre-NAC parameters |  |  |  |  |  |  |  |  |  |  |
| Age | [0 -50) | 678 | 210 | 1 |  |  | 0.716 |  |  |  |
|  | [50 -60) | 352 | 106 | 0.97 | [0.77 - 1.22] |  |  |  |  |  |
|  | 60+ | 167 | 55 | 1.11 | [0.82 - 1.49] |  |  |  |  |  |
| Menopausal status | Pre | 746 | 232 | 1 |  |  | 0.87 |  |  |  |
|  | Post | 442 | 135 | 0.98 | [0.79 - 1.21] |  |  |  |  |  |
| BMI | 18.5-24.9 | 680 | 193 | 1 |  |  | 0.009 | 1 |  |  |
|  | <18.5 | 48 | 15 | 1.13 | [0.67 - 1.91] | 0.651 |  | 1.13 | [ 0.66 - 1.91 ] | 0.66 |
|  | 25-29.9 | 304 | 96 | 1.15 | [0.9 - 1.47] | 0.255 |  | 1.06 | [ 0.83 - 1.36 ] | 0.624 |
|  | >=30 | 159 | 66 | 1.63 | [1.23 - 2.15] | <0.001 |  | 1.52 | [ 1.15 - 2.02 ] | 0.003 |
| Smoking status | No | 721 | 221 | 1 |  |  | 0.924 |  |  |  |
|  | Yes | 233 | 70 | 0.99 | [0.75 - 1.29] |  |  |  |  |  |
| BRCA mutation genes | BRCA1 | 31 | 9 | 1 |  |  | 0.991 |  |  |  |
|  | BRCA2 | 14 | 4 | 0.91 | [0.28 - 2.96] |  |  |  |  |  |
|  | others | 1 | 0 |  |  |  |  |  |  |  |
|  | No | 220 | 59 | 0.89 | [0.44 - 1.79] |  |  |  |  |  |
| Clinical T stage (TNM) | T0-T1 | 70 | 18 | 1 |  |  | <0.001 | 1 |  |  |
|  | T2 | 797 | 223 | 1.1 | [0.68 - 1.78] | 0.703 |  | 1.25 | [ 0.77 - 2.03 ] | 0.371 |
|  | T3-T4 | 329 | 129 | 1.79 | [1.09 - 2.93] | 0.021 |  | 1.69 | [ 1.02 - 2.78 ] | 0.04 |
| Clinical N stage (TNM) | N0 | 525 | 148 | 1 |  |  | 0.032 |  |  |  |
|  | N1-N2-N3 | 671 | 223 | 1.26 | [1.02 - 1.55] |  |  |  |  |  |
| BC subtype | Luminal | 526 | 184 | 1 |  |  | 0.025 | 1 |  |  |
|  | TNBC | 376 | 116 | 1.05 | [0.83 - 1.33] | 0.668 |  | 1.66 | [ 1.29 - 2.15 ] | <0.001 |
|  | HER2+ | 295 | 71 | 0.72 | [0.54 - 0.94] | 0.017 |  | 1.04 | [ 0.78 - 1.39 ] | 0.785 |
| ER status | Negative | 544 | 158 | 1 |  |  | 0.953 |  |  |  |
|  | Positive | 653 | 213 | 0.99 | [0.81 - 1.22] |  |  |  |  |  |
| PR status | Negative | 680 | 208 | 1 |  |  | 0.288 |  |  |  |
|  | Positive | 494 | 152 | 0.89 | [0.72 - 1.1] |  |  |  |  |  |
| Her2 status | Negative | 902 | 300 | 1 |  |  | 0.007 |  |  |  |
|  | Positive | 295 | 71 | 0.7 | [0.54 - 0.91] |  |  |  |  |  |
| Histological type | NST | 1060 | 317 | 1 |  |  | 0.106 |  |  |  |
|  | Others | 74 | 30 | 1.36 | [0.94 - 1.98] |  |  |  |  |  |
| KI67 | [0-10) | 65 | 21 | 1 |  |  | 0.494 |  |  |  |
|  | [10-20) | 110 | 38 | 1.07 | [0.63 - 1.82] |  |  |  |  |  |
|  | >=20 | 406 | 144 | 1.25 | [0.79 - 1.98] |  |  |  |  |  |
| SBR grade | Grade I-II | 477 | 170 | 1 |  |  | 0.11 |  |  |  |
|  | Grade III | 678 | 188 | 0.84 | [0.69 - 1.04] |  |  |  |  |  |
| **LVI** | No | 267 | 98 | 1 |  |  | 0.63 |  |  |  |
|  | Yes | 171 | 66 | 1.08 | [0.79 - 1.48] |  |  |  |  |  |
| **DCIS component** | No | 604 | 165 | 1 |  |  | 0.11 |  |  |  |
|  | Yes | 389 | 135 | 1.2 | [0.96 - 1.51] |  |  |  |  |  |
| **CT regimen (NAC)** | anthra-taxanes | 845 | 234 | 1 |  |  | **0.017** |  |  |  |
|  | anthra | 235 | 97 | 1.37 | [1.07 - 1.74] | **0.011** |  |  |  |  |
|  | taxanes | 25 | 4 | 0.59 | [0.22 - 1.59] | 0.3 |  |  |  |  |
|  | others | 91 | 36 | 1.42 | [1 - 2.02] | **0.052** |  |  |  |  |
| Post-NAC parameters |  |  |  |  |  |  |  |  |  |  |
| **pCR** | No pCR | 911 | 332 | 1 |  |  | **<0.001** |  |  |  |
|  | pCR | 285 | 39 | 0.35 | [0.25 - 0.49] | **<0.001** |  |  |  |  |
| **Post-NAC LVI** | No | 531 | 143 | 1 |  |  | **<0.001** |  |  |  |
|  | Yes | 302 | 144 | 2 | [1.59 - 2.52] | **<0.001** |  |  |  |  |
| **ypN** | 0 | 682 | 144 | 1 |  |  | **<0.001** | 1 | - | - |
|  | [1-3] | 341 | 127 | 1.8 | [1.42 - 2.28] | **<0.001** |  | 2.06 | [ 1.59 - 2.66 ] | **<0.001** |
|  | 4 and more | 174 | 100 | 3.35 | [2.59 - 4.32] | **<0.001** |  | 3.6 | [ 2.73 - 4.75 ] | **<0.001** |
| **RCB class** | RCB-0 | 202 | 23 | 1 |  |  | **<0.001** |  |  |  |
|  | RCB-I | 65 | 7 | 0.98 | [0.42 - 2.29] | 0.965 |  |  |  |  |
|  | RCB-II | 309 | 102 | 3.24 | [2.06 - 5.09] | **<0.001** |  |  |  |  |
|  | RCB-III | 141 | 72 | 5.56 | [3.47 - 8.89] | **<0.001** |  |  |  |  |

Abbreviations: pCR=pathological complete response; BMI=body mass index ; NST= no special type ; ER=estrogen receptor; PR=progesterone receptor; NAC=neoadjuvant chemotherapy; AC=anthracyclines; TILs=tumor infiltrating lymphocytes; RCB=residual cancer burden; LVI=lymphovascular invasion.

**Figure 3:**
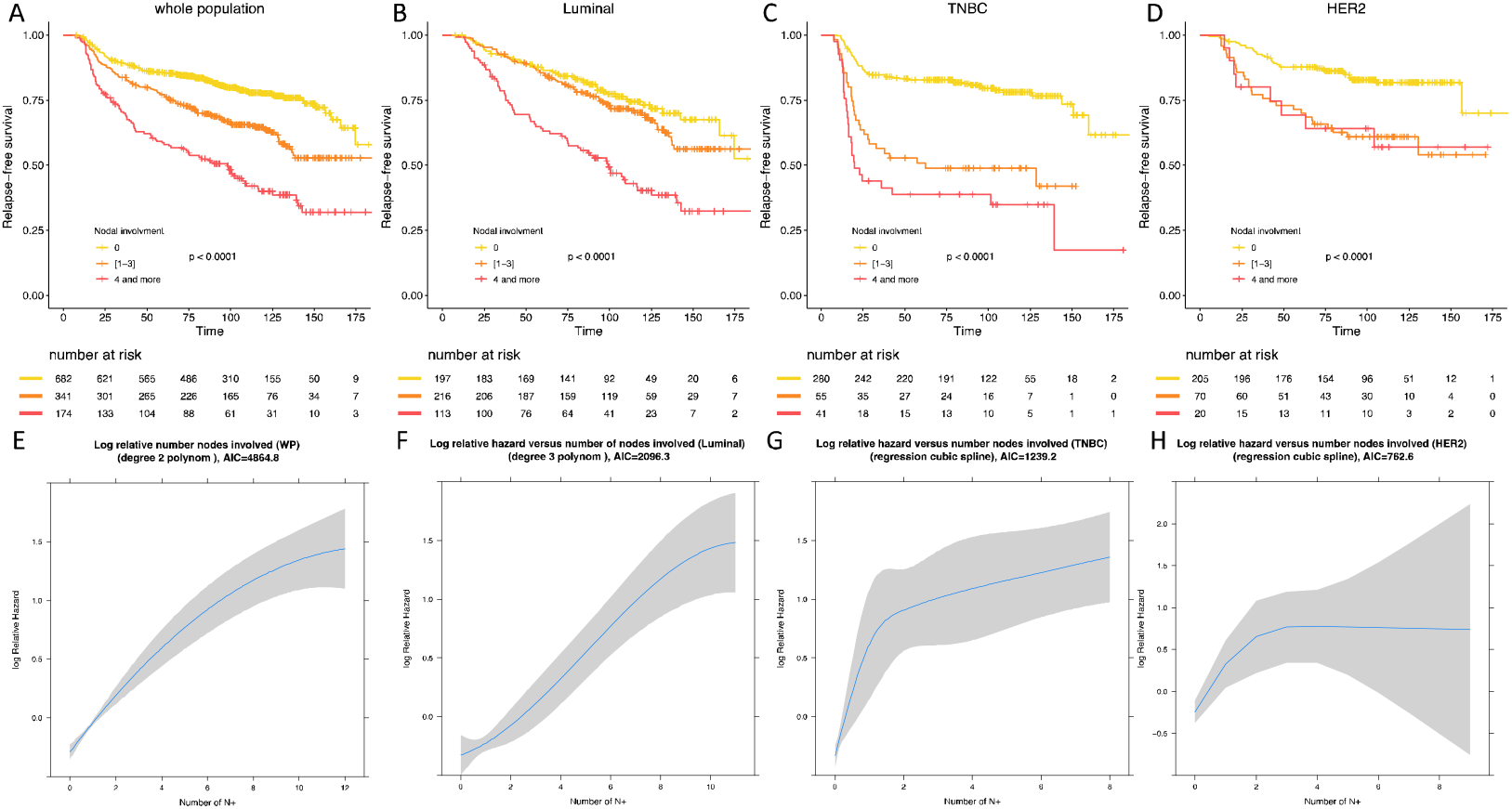
Relapse free survival according to BC subtype in the whole population (A), in luminal BC (B), in TNBC (C), in *HER-2* positive BC (D). Statistical models reflecting the association between relapse free survival and nodal status in the whole population (E), in luminal BC (F), in TNBC (G) and in *HER-2* positive BC (H).

**Figure 4:**
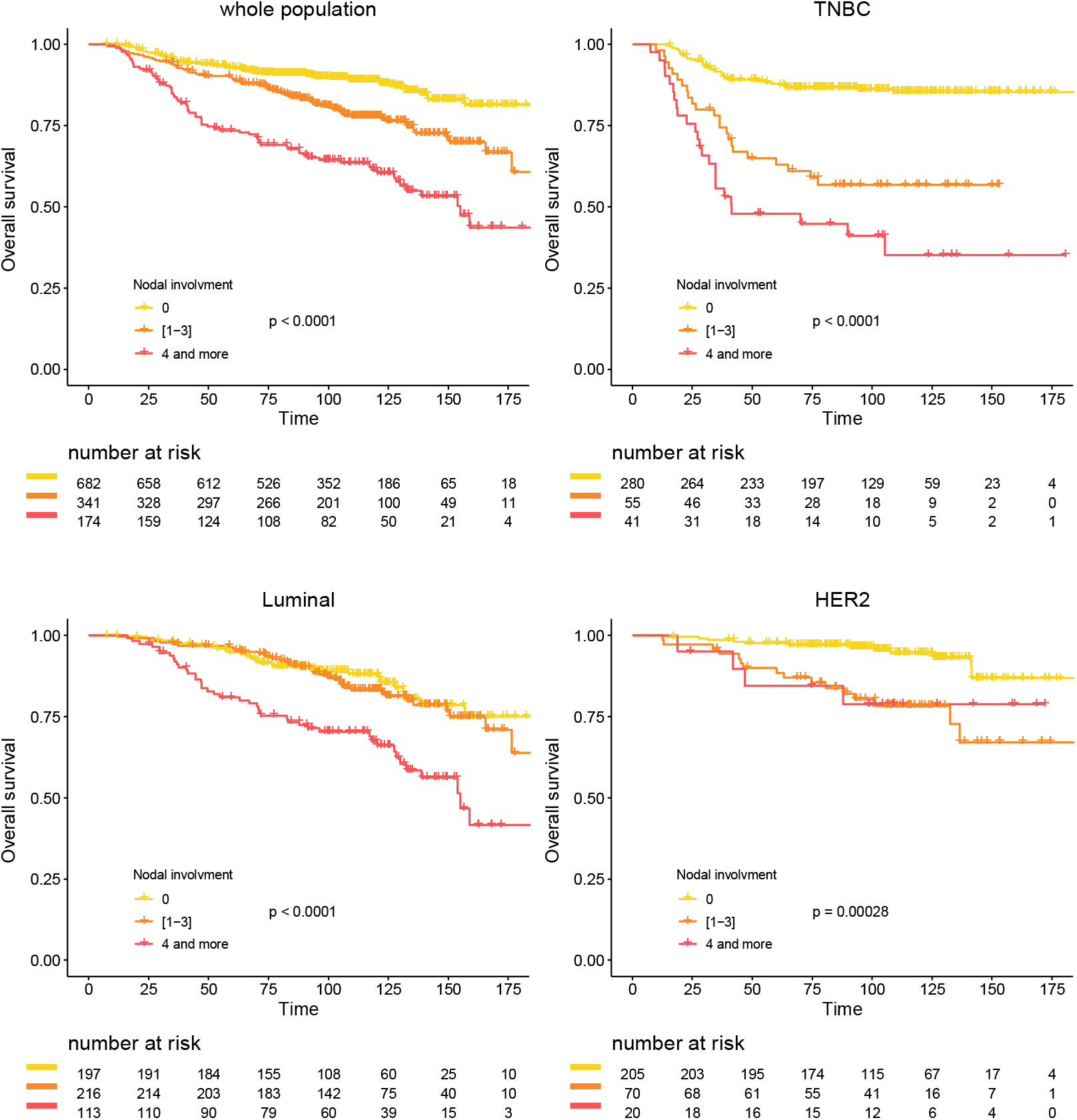
Overall survival according to post NAC nodal involvement in the whole population (A), Luminal BC (B), TNBC (C) and *HER-2* positive BC (D).

In luminal BCs, mild post-NAC nodal involvement was not associated with an impaired RFS when compared with ypN0 tumors (HR=1.24, 95%CI [0.86 - 1.79])(Table S1), whereas patients with a high nodal involvement were associated with an adverse prognosis (HR=2.8, 95%CI [1.93 - 4.06])(Fig3B). In TNBCs, both mild (HR=3.19, 95%CI [2.05 - 4.98])(Table S2) and high post-NAC nodal involvement (HR=4.83, 95%CI [3.06 - 7.63]) were associated with an impaired RFS when compared with ypN0 tumors. The difference between [1-3] and 4 and more was statistically significant (p<0.001) (Fig3C). In *HER2*-positive BCs, patients who had tumors with a mild nodal involvement were at a higher risk of relapse (HR=2.7, 95%CI [1.64 - 4.43])(Table S3) when compared with node negative tumors, but the prognosis was not significantly different from patients with 4 nodes involved or more (HR=2.69, 95%CI [1.24 - 5.8]) (Fig3D).

There was a significant deviation to the linearity assumption of the association between RFS and post-NAC nodal involvement in the whole population and in the 3 BC subtypes. After statistical modelisation, the statistical models best fitted a second-degree polynomial (whole population and luminal subgroup Fig 3E and 3F respectively), and a restricted cubic spline (TNBC and *HER2*-positive BCs, Fig 3G and 3H respectively).

After multivariate analysis (Table S1, S2, S3), post-NAC nodal involvement was significantly associated with RFS in luminal and TNBCs, but not in *HER2*-positive BC.

Similar results were obtained for overall survival (Figs 4A-D). The interaction between BC subtype, post-NAC nodal involvement and survival was highly significant (P_interaction_=0.005).

## Discussion

In this retrospective study of 1197 BC patients treated with NAC, we confirmed the strong prognostic value of nodal involvement after NAC, and we identified a marked difference in the prognostic impact of the axillar burden among the 3 BC subtypes.

Our study provides several new insights. First, it is in line with previous reports showing that the prognostic value of the axillary burden outperformed the value of the widely used binary endpoint pathological complete response. Rouzier *et al*. (12) found a higher correlation between RFS and axillary response to primary chemotherapy than with tumoral breast response in 152 BC patients. Hennessy *et al*. (13) found no impact of residual breast disease on survival outcomes among patients having achieved axillary pCR in a cohort of 403 BC patients with initial nodal involvement treated with NAC. This was confirmed by Dominici et al. (25) in 2010 in a retrospective study of 102 *HER-2* positive patients.

Second, along with previous studies (Table 4), we found a higher rate of post-NAC negative nodal status in case of TNBC, *HER2* positive BC, small tumor size, high grade tumors (10) (26) (27) (28) (29) (30) (31) (32) and high Ki67 (33) (34). In 2014, Boughey *et al*. (27) studied 694 BC patients treated with NAC with clinical nodal involvement at diagnosis. They found significantly higher rates of post-NAC ypN0 status among TNBC and *HER-2* positive BC subgroups (49.4% and 64.7% respectively) than in luminal BC patients (21.1%). In 2016, Mougalian et al. (10) found similar results in a cohort of 1600 stage II/III N+ BC patients: post-NAC negative nodal status rates repartition was 16.4% for luminal BC *versus* 40.8% for TNBCs and 47.3% for *HER-2* positive BC.

**Table 4:**
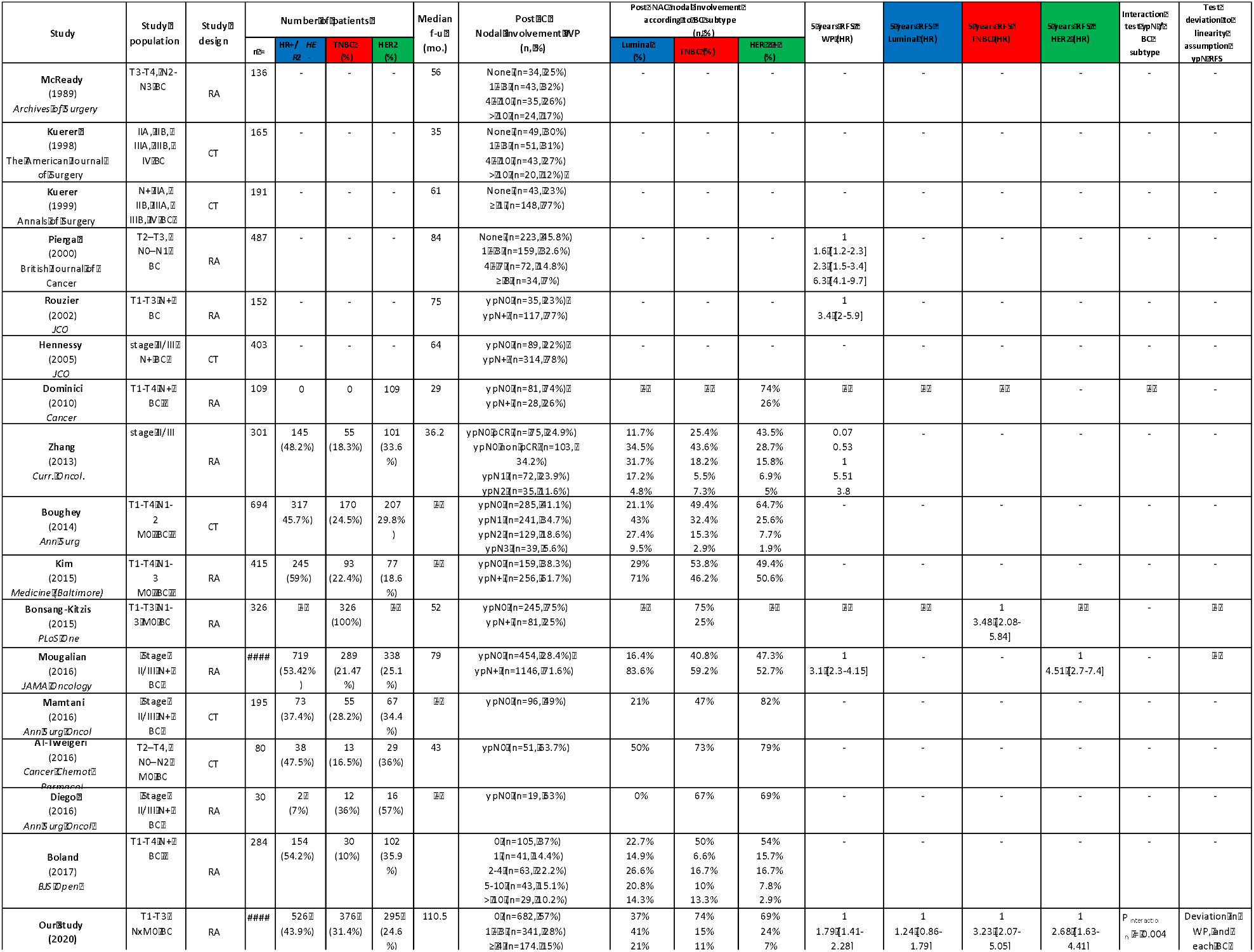
Summary of previous studies comparing prognosis according to nodal involvement after neoadjuvant chemotherapy (NAC) according to breast cancer subtype.

Little is known about the impact on survival outcome**s** of post NAC nodal involvement according to the number of positive nodes among BC subtypes. So far, most studies evaluating the prognostic impact of post-NAC nodal involvement used the binary endpoint ypN0 versus ypN+ (12) (10). Four studies ((9) (26) (27) (32)) used binned classes approaching the TNM classification (N0; N1: 1 to 3 nodes involved; N2: 4 to 9 nodes involved; N3: 10 or more nodes involved) (Table 4). However, to our knowledge, no study compared upfront the prognostic impact of nodal involvement according to BC subtypes nor performed linearity tests. Our results show that the prognostic value of the number of post-NAC positive nodes differs according to BC subtype. Patients with luminal BC presenting post-NAC axillary residual disease up to 3 positive nodes had a similar prognosis to those with no axillary residual disease, while we evidenced a negative impact on survival outcomes when the number of nodes involved was 4 or above. The prognostic impact of low-to-intermediate nodal involvement (1 to 3 nodes involved after chemotherapy) has also been studied in the adjuvant setting. Retrospective analyses from randomized trials have suggested that the recurrence score of a 21 gene assay (36) (37) could identify a subset of ER+/HER-2 negative BC patients with positive nodes who did not derive a significant benefit from chemotherapy: Albain *et al*. (38) and Dowsett *et al*. (39) found low risks of distant metastases in luminal low recurrence score N+ disease and luminal low recurrence score disease with 1 to 3 nodes involved respectively. The withholding of adjuvant chemotherapy for this category of BC patients is currently being evaluated in an ongoing trial (40). In the TNBC subgroup, as previously identified by our team(41), a positive nodal status after NAC was a poor prognostic factor, and the prognosis was worsened as soon as one lymph node was involved. However, as shown by the cubic spline statistical model best fitting the data, the slope of the increase of the risk was maximal between 0 and 2 lymph nodes, and the slope decreased thereafter. Finally, in the *HER-2* positive BC subgroup, the existence of residual axillary disease was a poor prognostic factor and the magnitude of the risk was similar for patients with 1 to 3 nodes involved and those with 4 or more nodes involved (RFS HR 2.68 95% CI [1.63-4.41] vs 2.67 95% CI [1.24-5.77]), though the interpretation might be limited by the weak effective of the latter category (6.8% of patients with *HER-2* positive BC).

To the best of our knowledge, we report here the first upfront comparison of the prognostic value of residual axillary disease among each BC subtype, while taking into account the number of positive nodes after NAC. In addition, we evidenced that the relationship between nodal involvement and relapse free survival was nonlinear, and this was true in every BC subtype. The main strengths of our study include its large statistical power, its long-term follow-up. Limits of our study include its retrospective design and the absence of external independent validation.

Our study has pragmatic implications. If confirmed in independent studies, it suggests that the cut-off to consider high risk patients after NAC completion should be different according to BC subtypes: 4 or more nodes involved for luminal BC patients, and 1 for TNBC and *HER- 2* positive BC patients. With the widespread routine use of NAC for TNBC and *HER2*- positive BC patients (1) (2), second-line trials in the post neoadjuvant setting for high risk patients are increasing(42) testing the addition of chemotherapy (ECOG-ACRIN Cancer Research phase III trial in TNBC NCT02445391), PARP inhibitors (phase III OLYMPIA trial in *HER-2* negative BC NCT02032823), immunotherapy (Nab-Paclitaxel and Atezolizumab in TNBC NCT02530489; Pembrolizumab in TNBC NCT02954874; Avelumab in TNBC NCT02926196), cycline-dependent kinase inhibitors (phase III PENELOPE B study for HR positive BC NCT01864746) or vaccines (nelipepimut-S/GM-CSF in *HER-2* positive BC NCT02297698; WOKVAC and DC1 in *HER-2* positive BC NCT03384914). Our findings are of particular importance since they may help to identify more accurately high-risk patients who might benefit from such treatments by considering the number of residual positive nodes after NAC as a cornerstone of prognostication.

## Data Availability

All data are available

## Acknowledgements

We thank Roche France for financial support for the construction of the Institut Curie neoadjuvant database (NEOREP).

