## Supplemental material for "The prognostic value of lymph node involvement after neoadjuvant chemotherapy is different among breast cancer subtypes"

**SUPPLEMENTAL MATERIAL, TABLES AND FIGURES**

**Supplementary Material:**

***Pathological review:***

*Residual Cancer Burden*

To enable the calculation of RCB score, the following variables were retrieved: diameter of the primary tumor bed in the resection specimen ($d_{prim}$), the proportion of the primary tumor bed that contained invasive carcinoma ($f_{inv}$), the number of axillary lymph nodes containing metastatic carcinoma (LN), and the diameter of the largest metastasis in an axillary lymph node ($d_{met}$). Largest bidimensional measurements of the residual primary tumor bed were recorded from the macroscopic description in the pathology report. If multiple tumors were present, the dimensions of the largest were considered. The proportion of invasive carcinoma (finv) within the cross sectional area of the primary tumor bed was estimated from the overall percent area of carcinoma (%CA) and then corrected for the component of in situ carcinoma (%CIS): $f_{inv}$ (1 – (%CIS/100)) (%CA/100).

RCB was calculated with the formule: $RCB = 1.4( f_{inv}\ d_{prim})^{0.17} + [4 (1 - 0.75^{LN})\ d_{met}]^{0.17}$

*Lymphovascular invasion*

Presence or absence of LVI was determined by unstained standard formalin-fixed paraffin-embedded examination on surgical specimen. Immunostaining with vascular markers was occasionally performed to rule out invasive carcinoma with shrinkage artifact. LVI data were extracted from pathology records by two independent researchers (TL, ASH), and were dichotomized into a binary variable (Post-NAC LVI: yes/ no). Patients whose tumor reached pCR were considered as having no LVI. Results were crosschecked and a breast expert pathologist (ML) resolved discrepancies.

*Tumor infiltrating lymphocytes*

In accordance with the recommendations of the international TILs Working Group (Dieci et al., 2018), we checked for presence of a mononuclear cell infiltrate in the stroma on hematoxylin and eosin-stained sections without additional staining, after excluding areas around ductal carcinomas in situ (DCIS), and tumor zones with necrosis and artifacts. Infiltrates were scored on a continuous scale, as the mean percentage of the stromal area occupied by mononuclear cells. After NAC, we assessed TIL levels within the borders of the residual tumor bed, as defined by the RCB index (Symmans et al., 2007b). In cases of pCR, the scar area was measured on macroscopic examination. The scar appeared as a white area in the breast parenchyma corresponding to the tumor bed modified by NAC. It was characterized by the presence of histiocytes, lymphocytes, macrophages, fibrosis and elastosis. The whole fibro-inflammatory scar was evaluated on HE sections (size in mm and stromal TIL level evaluation)

**Supplementary Tables:**

**Supplementary Table 1: Association between clinical and pathological pre and post-NAC parameters with overall-free survival (Luminal BC population, univariate and multivariate analysis).**

*p* represents the p-value for the Wald test, and p* represents the individual p-value versus reference class. Abbreviations: pCR=pathological complete response; BMI=body mass index ; NST= no special type ; ER=estrogen receptor; PR=progesterone receptor; NAC=neoadjuvant chemotherapy; AC=anthracyclines; TILs=tumor infiltrating lymphocytes; RCB=residual cancer burden; LVI=lymphovascular invasion.

**Supplementary Table 2: Association between clinical and pathological pre and post-NAC parameters with overall-free survival (TNBC population, univariate and multivariate analysis).**

*p* represents the p-value for the Wald test, and p* represents the individual p-value versus reference class. Abbreviations: pCR=pathological complete response; BMI=body mass index ; NST= no special type ; ER=estrogen receptor; PR=progesterone receptor; NAC=neoadjuvant chemotherapy; AC=anthracyclines; TILs=tumor infiltrating lymphocytes; RCB=residual cancer burden; LVI=lymphovascular invasion.

**Supplementary Table 3: Association between clinical and pathological pre and post-NAC parameters with overall-free survival (*HER-2* BC population, univariate and multivariate analysis).**

*p* represents the p-value for the Wald test, and p* represents the individual p-value versus reference class. Abbreviations: pCR=pathological complete response; BMI=body mass index

; NST= no special type ; ER=estrogen receptor; PR=progesterone receptor; NAC=neoadjuvant chemotherapy; AC=anthracyclines; TILs=tumor infiltrating lymphocytes; RCB=residual cancer burden; LVI=lymphovascular invasion.

**Supplementary Table 1: Association between clinical and pathological pre and post-NAC parameters with overall-free survival (Luminal BC population, univariate and multivariate analysis).**

*p* represents the p-value for the Wald test, and p* represents the individual p-value versus reference class. Abbreviations: pCR=pathological complete response; BMI=body mass index ; NST= no special type ; ER=estrogen receptor; PR=progesterone receptor; NAC=neoadjuvant chemotherapy; AC=anthracyclines; TILs=tumor infiltrating lymphocytes; RCB=residual cancer burden; LVI=lymphovascular invasion.

| Luminal | | | | | | | | | | |
|---|---|---|---|---|---|---|---|---|---|---|
| | | | | Univariate | | | | Multivariate | | |
| **Variable** | **Class** | **Number** | **Events** | **HR** | **CI** | **p*** | ***p*** | **HR** | **CI** | **p** |
| Pre-NAC parameters | | | | | | | | | | |
| **Age** | [0 -50) | 304 | 101 | 1 | | | 0.345 | | | |
| | [50 -60) | 158 | 56 | 1.06 | [0.76 - 1.46] | | | | | |
| | 60+ | 64 | 27 | 1.37 | [0.9 - 2.1] | | | | | |
| **Menopausal status** | Pre | 338 | 111 | 1 | | | 0.212 | | | |
| | Post | 184 | 71 | 1.21 | [0.9 - 1.63] | | | | | |
| **BMI** | 18.5-24.9 | 289 | 91 | 1 | | | **0.053** | | | |
| | <18.5 | 24 | 10 | 1.63 | [0.85 - 3.14] | 0.141 | | | | |
| | 25-29.9 | 135 | 46 | 1.08 | [0.76 - 1.54] | 0.682 | | | | |
| | >=30 | 75 | 36 | 1.64 | [1.11 - 2.41] | **0.012** | | | | |
| **Smoking status** | No | 297 | 107 | 1 | | | 0.952 | | | |
| | Yes | 113 | 38 | 1.01 | [0.7 - 1.46] | | | | | |
| **BRCA mutation genes** | BRCA1 | 8 | 2 | 1 | | | 0.997 | | | |
| | BRCA2 | 6 | 2 | 1.24 | [0.17 - 8.82] | | | | | |
| | others | 1 | 0 | | | | | | | |
| | No | 74 | 24 | 1.16 | [0.27 - 4.91] | | | | | |
| **Clinical T stage (TNM)** | T0-T1 | 21 | 10 | 1 | | | **0.01** | | | |
| | T2 | 358 | 111 | 0.59 | [0.31 - 1.14] | 0.115 | | | | |
| | T3-T4 | 147 | 63 | 0.93 | [0.48 - 1.81] | 0.824 | | | | |
| **Clinical N stage (TNM)** | N0 | 235 | 75 | 1 | | | 0.1 | | | |
| | N1-N2-N3 | 290 | 109 | 1.28 | [0.95 - 1.72] | | | | | |
| **Histological type** | NST | 444 | 150 | 1 | | | 0.08 | | | |
| | Others | 65 | 28 | 1.43 | [0.96 - 2.15] | | | | | |
| **KI67** | [0-10) | 52 | 17 | 1 | | | 0.778 | | | |
| | [10-20) | 79 | 29 | 1.13 | [0.62 - 2.05] | | | | | |
| | >=20 | 157 | 58 | 1.21 | [0.71 - 2.08] | | | | | |
| **SBR grade** | Grade I-II | 324 | 119 | 1 | | | 0.643 | | | |
| | Grade III | 182 | 58 | 0.93 | [0.68 - 1.27] | | | | | |
| **LVI** | No | 144 | 59 | 1 | | | 0.162 | | | |

| | | | | | | | | | | |
|---|---|---|---|---|---|---|---|---|---|---|
| | Yes | 84 | 27 | 0.72 | [0.46 - 1.14] | | | | | |
| **DCIS component** | No | 186 | 59 | 1 | | | 0.677 | | | |
| | Yes | 224 | 81 | 1.07 | [0.77 - 1.51] | | | | | |
| **CT regimen (NAC)** | anthra-taxans | 340 | 108 | 1 | | | 0.272 | | | |
| | anthra | 133 | 54 | 1.05 | [0.75 - 1.46] | | | | | |
| | taxanes | 8 | 1 | 0.37 | [0.05 - 2.64] | | | | | |
| | others | 45 | 21 | 1.49 | [0.93 - 2.38] | | | | | |
| Post-NAC parameters | | | | | | | | | | |
| **pCR** | No pCR | 493 | 179 | 1 | | | **0.09** | | | |
| | pCR | 33 | 5 | 0.46 | [0.19 - 1.13] | **0.09** | | | | |
| **Post-NAC LVI** | No | 237 | 75 | 1 | | | **0.046** | | | |
| | Yes | 189 | 78 | 1.38 | [1.01 - 1.9] | **0.046** | | | | |
| **ypN** | 0 | 197 | 49 | 1 | | | **<0.001** | 1 | | |
| | [1-3] | 216 | 70 | 1.24 | [0.86 - 1.79] | 0.245 | | 1.18 | [ 0.82 - 1.71 ] | 0.377 |
| | 4 and more | 113 | 65 | 2.8 | [1.93 - 4.06] | **<0.001** | | 2.68 | [ 1.84 - 3.89 ] | **<0.001** |
| **RCB class** | RCB-0 | 11 | 3 | 1 | | | 0.172 | | | |
| | RCB-I | 18 | 2 | 0.41 | [0.07 - 2.43] | | | | | |
| | RCB-II | 109 | 33 | 1.12 | [0.34 - 3.64] | | | | | |
| | RCB-III | 84 | 36 | 1.58 | [0.48 - 5.13] | | | | | |

**Supplementary Table 2: Association between clinical and pathological pre and post-NAC parameters with overall-free survival (TNBC population, univariate and multivariate analysis).**

*p* represents the p-value for the Wald test, and p* represents the individual p-value versus reference class. Abbreviations: pCR=pathological complete response; BMI=body mass index ; NST= no special type ; ER=estrogen receptor; PR=progesterone receptor; NAC=neoadjuvant chemotherapy; AC=anthracyclines; TILs=tumor infiltrating lymphocytes; RCB=residual cancer burden; LVI=lymphovascular invasion.

| TNBC | | | | | | | | | | |
|---|---|---|---|---|---|---|---|---|---|---|
| | | | | Univariate | | | | Multivariate | | |
| **Variable** | **Class** | **Number** | **Events** | **HR** | **CI** | **p*** | ***p*** | **HR** | **CI** | **p** |
| Pre-NAC parameters | | | | | | | | | | |
| **Age** | [0 -50) | 205 | 60 | 1 | | | 0.723 | | | |
| | [50 -60) | 115 | 38 | 1.17 | [0.78 - 1.77] | | | | | |
| | 60+ | 56 | 18 | 1.13 | [0.67 - 1.91] | | | | | |
| **Menopausal status** | Pre | 229 | 70 | 1 | | | 0.991 | | | |
| | Post | 143 | 44 | 1 | [0.69 - 1.46] | | | | | |
| **BMI** | 18.5-24.9 | 211 | 65 | 1 | | | 0.238 | | | |
| | <18.5 | 12 | 1 | 0.22 | [0.03 - 1.58] | | | | | |
| | 25-29.9 | 103 | 31 | 1.01 | [0.66 - 1.54] | | | | | |
| | >=30 | 49 | 19 | 1.4 | [0.84 - 2.33] | | | | | |
| **Smoking status** | No | 238 | 73 | 1 | | | 0.565 | | | |
| | Yes | 66 | 18 | 0.86 | [0.51 - 1.44] | | | | | |
| **BRCA mutation genes** | BRCA1 | 22 | 6 | 1 | | | 0.673 | | | |
| | BRCA2 | 5 | 2 | 1.43 | [0.29 - 7.1] | | | | | |
| | No | 83 | 18 | 0.79 | [0.31 - 1.98] | | | | | |
| **Clinical T stage (TNM)** | T0-T1 | 31 | 7 | 1 | | | **0.001** | 1 | | |
| | T2 | 243 | 65 | 1.23 | [0.56 - 2.69] | 0.6 | | 1.28 | [0.58 - 2.8] | 0.544 |
| | T3-T4 | 102 | 44 | 2.42 | [1.09 - 5.38] | **0.03** | | 2.29 | [1.03 - 5.09] | **0.043** |
| **Clinical N stage (TNM)** | N0 | 171 | 49 | 1 | | | 0.282 | | | |
| | N1-N2-N3 | 205 | 67 | 1.22 | [0.85 - 1.77] | | | | | |
| **Histological type** | NST | 340 | 101 | 1 | | | 0.367 | | | |
| | Others | 6 | 1 | 0.4 | [0.06 - 2.9] | | | | | |
| **KI67** | [0-10) | 10 | 2 | 1 | | | 0.558 | | | |
| | [10-20) | 16 | 6 | 2.18 | [0.44 - 10.81] | | | | | |
| | >=20 | 141 | 52 | 2.18 | [0.53 - 8.93] | | | | | |
| **SBR grade** | Grade I-II | 53 | 17 | 1 | | | 0.917 | | | |
| | Grade III | 314 | 97 | 1.03 | [0.61 - 1.72] | | | | | |
| **LVI** | No | 82 | 28 | 1 | | | 0.121 | | | |
| | Yes | 38 | 18 | 1.6 | [0.88 - 2.9] | | | | | |

| | | | | | | | | | | |
|---|---|---|---|---|---|---|---|---|---|---|
| **DCIS component** | No | 282 | 86 | 1 | | | 0.219 | | | |
| | Yes | 51 | 21 | 1.35 | [0.84 - 2.18] | | | | | |
| **CT regimen (NAC)** | anthra-taxanes | 287 | 86 | 1 | | | 0.728 | | | |
| | anthra | 63 | 24 | 1.2 | [0.76 - 1.89] | | | | | |
| | taxanes | 1 | 0 | | | | | | | |
| | others | 24 | 6 | 0.73 | [0.32 - 1.69] | | | | | |
| Post-NAC parameters | | | | | | | | | | |
| **pCR** | No pCR | 236 | 96 | 1 | | | <0.001 | | | |
| | pCR | 139 | 20 | 0.29 | [0.18 - 0.47] | <0.001 | | | | |
| **Post-NAC LVI** | No | 170 | 48 | 1 | | | <0.001 | | | |
| | Yes | 64 | 38 | 2.65 | [1.73 - 4.06] | <0.001 | | | | |
| **ypN** | 0 | 280 | 60 | 1 | | | <0.001 | 1 | | |
| | [1-3] | 55 | 29 | 3.19 | [2.05 - 4.98] | <0.001 | | 3.17 | [2.03 - 4.95] | <0.001 |
| | 4 and more | 41 | 27 | 4.83 | [3.06 - 7.63] | <0.001 | | 4.52 | [2.85 - 7.17] | <0.001 |
| **RCB class** | RCB-0 | 123 | 18 | 1 | | | <0.001 | | | |
| | RCB-I | 23 | 4 | 1.22 | [0.41 - 3.61] | 0.717 | | | | |
| | RCB-II | 131 | 50 | 3.09 | [1.8 - 5.29] | <0.001 | | | | |
| | RCB-III | 42 | 30 | 9.16 | [5.08 - 16.54] | <0.001 | | | | |

**Supplementary Table 3: Association between clinical and pathological pre and post-NAC parameters with overall-free survival (*HER-2* BC population, univariate and multivariate analysis).**

*p* represents the p-value for the Wald test, and p* represents the individual p-value versus reference class. Abbreviations: pCR=pathological complete response; BMI=body mass index ; NST= no special type ; ER=estrogen receptor; PR=progesterone receptor; NAC=neoadjuvant chemotherapy; AC=anthracyclines; TILs=tumor infiltrating lymphocytes; RCB=residual cancer burden; LVI=lymphovascular invasion.

| *HER2*-positive | | | | | | | | | | |
|---|---|---|---|---|---|---|---|---|---|---|
| | | | | Univariate | | | | Multivariate | | |
| **Variable** | **Class** | **Number** | **Events** | **HR** | **CI** | **p*** | ***p*** | **HR** | **CI** | **p** |
| Pre-NAC parameters | | | | | | | | | | |
| **Age** | [0 -50) | 169 | 49 | 1 | | | 0.066 | | | |
| | [50 -60) | 79 | 12 | 0.48 | [0.25 - 0.9] | **0.022** | | | | |
| | 60+ | 47 | 10 | 0.74 | [0.37 - 1.46] | 0.386 | | | | |
| **Menopausal status** | Pre | 179 | 51 | 1 | | | **0.039** | 1 | | |
| | Post | 115 | 20 | 0.58 | [0.35 - 0.97] | **0.039** | | 0.44 | [ 0.23 - 0.84 ] | **0.013** |
| **BMI** | 18.5-24.9 | 180 | 37 | 1 | | | 0.253 | | | |
| | <18.5 | 12 | 4 | 1.74 | [0.62 - 4.89] | | | | | |
| | 25-29.9 | 66 | 19 | 1.52 | [0.87 - 2.64] | | | | | |
| | >=30 | 35 | 11 | 1.71 | [0.87 - 3.37] | | | | | |
| **Smoking status** | No | 186 | 41 | 1 | | | 0.563 | | | |
| | Yes | 54 | 14 | 1.2 | [0.65 - 2.2] | | | | | |
| **BRCA mutation genes** | BRCA1 | 1 | 1 | 1 | | | 0.423 | | | |
| | BRCA2 | 3 | 0 | 0 | [0 - Inf] | | | | | |
| | No | 63 | 17 | 0.26 | [0.03 - 1.96] | | | | | |
| **Clinical T stage (TNM)** | T0-T1 | 18 | 1 | 1 | | | 0.167 | | | |
| | T2 | 196 | 47 | 4.69 | [0.65 - 34] | 0.126 | | | | |
| | T3-T4 | 80 | 22 | 6.06 | [0.82 - 44.95] | 0.078 | | | | |
| **Clinical N stage (TNM)** | N0 | 119 | 24 | 1 | | | 0.208 | | | |
| | N1-N2-N3 | 176 | 47 | 1.37 | [0.84 - 2.24] | | | | | |
| **Histological type** | NST | 276 | 66 | 1 | | | 0.645 | | | |
| | Others | 3 | 1 | 1.59 | [0.22 - 11.48] | | | | | |
| **KI67** | [0-10) | 3 | 2 | 1 | | | 0.244 | | | |
| | [10-20) | 15 | 3 | 0.22 | [0.04 - 1.3] | | | | | |
| | >=20 | 108 | 34 | 0.42 | [0.1 - 1.76] | | | | | |
| **SBR grade** | Grade I-II | 100 | 34 | 1 | | | **0.006** | 1 | | |
| | Grade III | 182 | 33 | 0.51 | [0.32 - 0.83] | **0.006** | | 0.39 | [ 0.23 - 0.68 ] | **0.001** |
| **LVI** | No | 41 | 11 | 1 | | | 0.085 | | | |
| | Yes | 49 | 21 | 1.91 | [0.91 - 3.99] | 0.085 | | | | |
| **DCIS component** | No | 136 | 20 | 1 | | | **0.006** | | | |

| | | | | | | | |
|---|---|---|---|---|---|---|---|
| | Yes | 114 | 33 | 2.19 | [1.25 - 3.82] | **0.006** | |
| **CT regimen (NAC)** | anthra-taxanes | 218 | 40 | 1 | | | **<0.001** |
| | anthra | 39 | 19 | 3.2 | [1.85 - 5.54] | **<0.001** | |
| | taxanes | 16 | 3 | 1.12 | [0.35 - 3.63] | 0.848 | |
| | others | 22 | 9 | 2.64 | [1.28 - 5.45] | **0.009** | |
| Post-NAC parameters | | | | | | | |
| **pCR** | No pCR | 182 | 57 | 1 | | | **<0.001** |
| | pCR | 113 | 14 | 0.35 | [0.2 - 0.64] | **<0.001** | |
| **Post-NAC LVI** | No | 124 | 20 | 1 | | | **<0.001** |
| | Yes | 49 | 28 | 4.93 | [2.77 - 8.78] | **<0.001** | |
| **ypN** | 0 | 205 | 35 | 1 | | | **<0.001** |
| | [1-3] | 70 | 28 | 2.7 | [1.64 - 4.43] | **<0.001** | |
| | 4 and more | 20 | 8 | 2.69 | [1.24 - 5.8] | **0.012** | |
| **RCB class** | RCB-0 | 68 | 2 | 1 | | | **0.002** |
| | RCB-I | 24 | 1 | 1.52 | [0.14 - 16.78] | 0.732 | |
| | RCB-II | 69 | 19 | 10.35 | [2.41 - 44.46] | **0.002** | |
| | RCB-III | 15 | 6 | 16.53 | [3.33 - 81.99] | **<0.001** | |

**Supplementary Figures:**

**Supplementary Figure 1:** Association between post-NAC involvement and tumor characteristics according to BC subtype: RCB index (A); Lymphovascular invasion (B); Tumor cellularity (C); Post-NAC mitotic index (D); Stromal TIL levels (E). Intra tumoral (IT) TIL levels (F).

**Supplementary Figure 1:** Association between post-NAC involvement and tumor characteristics according to BC subtype: RCB index (A); Lymphovascular invasion (B); Tumor cellularity (C); Post-NAC mitotic index (D); Stromal TIL levels (E). Intra tumoral (IT) TIL levels (F).

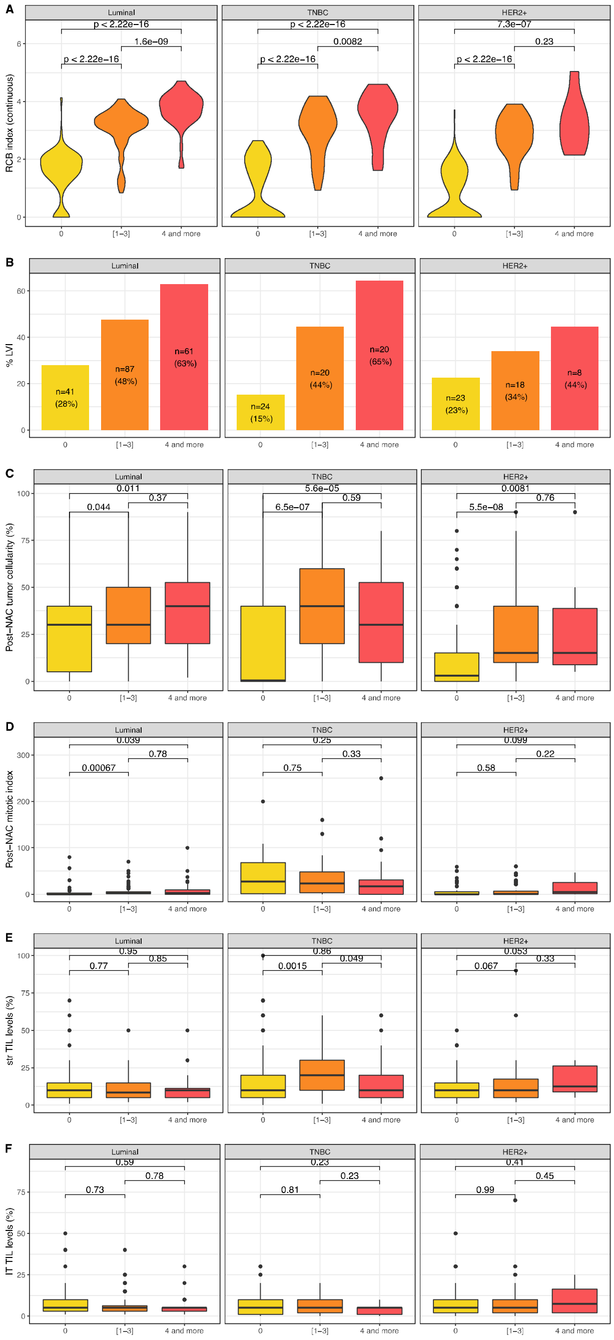
